# Clinician-Trained Artificial Intelligence for Enhanced Routing of Patient Portal Messages in the Electronic Health Record

**DOI:** 10.1101/2023.11.27.23298910

**Authors:** Arash Harzand, Muhammad Zia ul Haq, Andrew M. Hornback, Alison D. Cowan, Blake Anderson

## Abstract

**Background:** Increased use of electronic health records (EHR) and patient-initiated messaging has led to inefficiency, staff shortages, and provider burnout. We evaluated the impact of a natural language processing (NLP) algorithm for message routing on communication dynamics.

**Methods:** We developed an NLP model to accurately label inbound EHR messages from patients using a pre-trained classifier with fine-tuning based on clinician feedback. In a real-world study, the model was deployed to prospectively label and route messages sent to participating physicians at an integrated health system. A parallel control group of unrouted messages was generated for comparative analysis. The primary endpoints for assessing model performance were the time to first message interaction, the time to conversation resolution, and the total number of message interactions by staff, compared with the control group. Secondary endpoints were the precision, recall, F1 score (a measure of positive predictive value and sensitivity), and accuracy for correct message classification.

**Results:** The model prospectively labeled and routed 469 unique conversations over 14 days from the inbaskets of participating physicians. Compared to a control group of 402 unrouted conversations from the same time period, conversations in the routed group had an 83.3% reduction in the time to first interaction (median difference [MD], −1 hour; P<0.001), an 84.4% reduction in the time to conversation resolution (MD, −22.5 hours; P<0.001), and a 40% reduction in the total number of staff interactions after application of intervention (MD, −2.0 interactions; P<0.001). The model demonstrated high precision (>97.6%), recall (>95%), and F1 scores (>96.5%) for accurate prediction of all five message classes, with a total accuracy of 97.8%.

**Conclusions:** Real-time message routing using advanced NLP was associated with significantly reduced message response and resolution times.

## INTRODUCTION

The electronic health record (EHR) has become the primary documentation mechanism for healthcare data during the last decade.^1^ In the United States, healthcare policies such as the CMS Meaningful Use Program have incentivized rapid EHR adoption to enhance healthcare quality.^2–4^ Conversely, healthcare staff burnout has increased in parallel and is largely attributed to the increased time spent performing administrative tasks in the EHR. The EHR is cited as a primary source of physician burnout, affecting seven out of 10 physicians.^5,6^ Physicians spend twice the amount on EHR-related tasks such as documentation and messaging for each 1 hour spent providing direct patient care.^7^

Studies have shown that patients increasingly use EHR messaging portals as an alternative to seeking in-person care due to the COVID-19 pandemic.^8–10^ In many clinical practices, these patient-initiated messages (PIMs) are directed first to a central pool before being directed to a target staff member to either act on a message or who may forward it to another user. Successive forwarding and replying can result in exponential growth of staff messaging interactions, with each “touch” representing a new cognitive load for the interactor, distraction from other tasks, or additional time spent after the workday. An initial PIM has been shown to generate as many as seven or more staff touches before final resolution.

The increased workload from PIMs has led to calls for novel methods to enhance the routing and management of EHR-based messaging. Prior approaches to reduce provider burden have included efforts at EHR inbox redesign, team-based support, and dedicated triage teams.^11–13^ Deploying natural language processing (NLP) is an emerging approach for addressing burnout and reducing cognitive overload. We have recently demonstrated the feasibility of using NLP to automatically prioritize urgent patient messages and identify public health trends from the EHR.^14^ Although additional studies have also shown the potential of using text search and NLP to analyze and label messages effectively,^15–19^ none have explored the value of adding machine learning (ML) to automate message triage through intelligent rerouting.^20^ No previously published efforts have used high-level NLP to analyze and intelligently reroute patient messages.

The present study aimed to develop and validate an ML classifier for intelligent routing of inbound EHR messages and measure performance in a real-world clinical setting. The objectives were to 1) create a gold standard classifier using manual message validation by multi-specialty physicians and nurses, 2) refine and validate an NLP classifier to accurately label messages based on common message themes (e.g., scheduling requests or refills), and 3) to evaluate the model’s ability to reduce the time and amount of message interactions required by staff for message resolution.

## METHODS

### Study Design, Population, and Data Sources

This prospective, real-world study was conducted across four outpatient locations at Emory Healthcare, an integrated health system in Atlanta, Georgia (**Figure 1)**. Individual physicians were invited to participate and have their inbound EHR message pools included in the NLP intervention. All messages from adult patients (aged ≥18 years) for each participating physician were accessed from the EHR (PowerChart EHR, Oracle Cerner, Kansas City, MO) and included in the intervention group. An observational cohort drawn randomly from unrouted messages from the same time period was generated for comparative analysis. The study was deemed exempt from human subjects research by the Emory University Institutional Review Board and received a waiver of informed consent based on the no more than minimal risk to patients.

**Figure 1.**
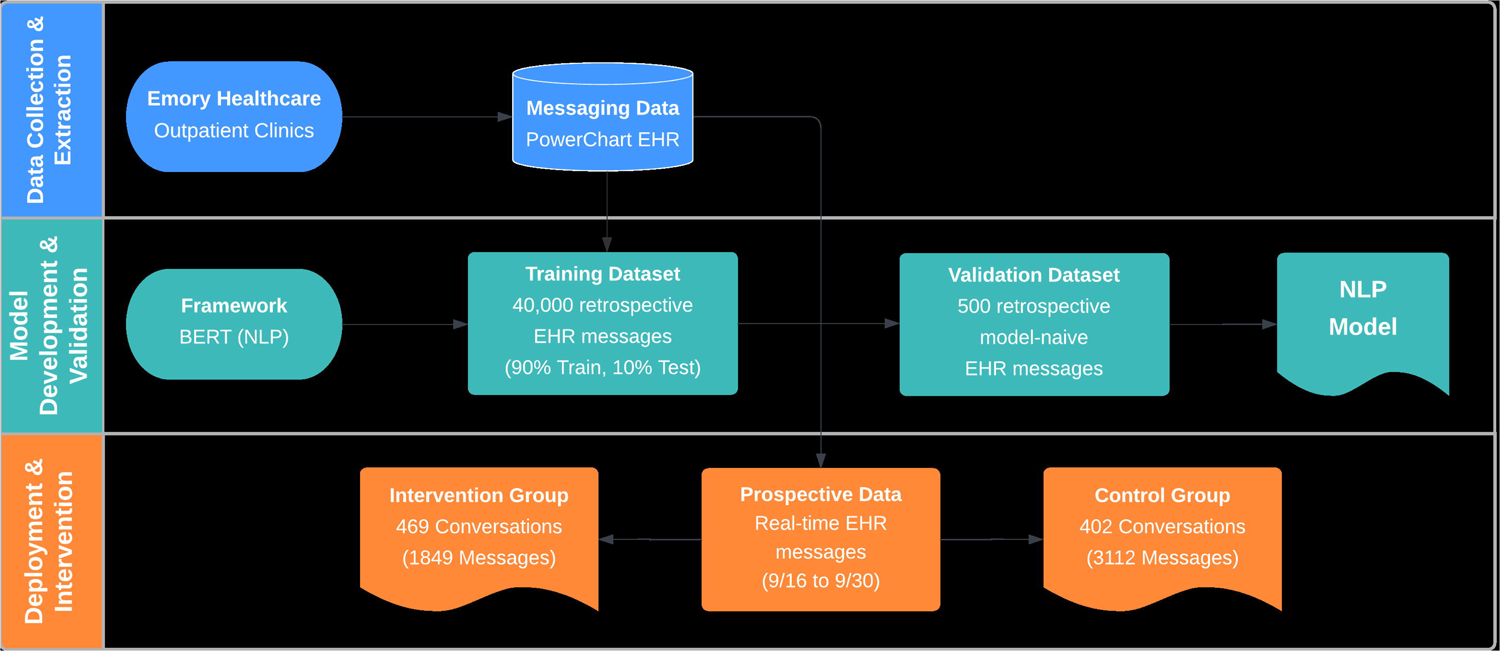
Data extraction, model development, and intervention flowchart

### Model Development and Validation

We trained an NLP model using the Bidirectional Encoder Representations from Transformers (BERT) language representation model.^21^ The model’s primary objective was to correctly label inbound messages and route them to the appropriate staff based on the predicted message classification. To develop the model, we randomly sampled a retrospective corpus of 40,000 EHR messages from adult patients that were expertly labeled by a team of study clinicians (four physicians; one nurse) into one of five message classes: “urgent”, “clinician”, “refill”, “schedule”, or “form”. We then randomly divided the labeled dataset into training (90%) and testing (10%) cohorts which were used to fine-tune the pre-trained BERT model. For post-training validation, the study clinicians reviewed the model’s classification accuracy using a sequestered set of 500 model-naive messages.

### Study Intervention and Model Deployment

To evaluate real-world performance, the fully trained and validated model was prospectively deployed to EHR messaging pools for all participating physicians to monitor and route all incoming messages from adult patients. The model was designed to operate silently within the EHR backend to minimize added workflow steps for staff and enable model evaluation under real-world conditions. Prospective model performance was measured and reported separately for each class using a correlation heat map and confusion matrix (**Figure 3**). The precision, recall, F1 score, and accuracy were also calculated for each class to determine the model’s ability to correctly classify instances of each class, its sensitivity to the instances of the class, the harmonic mean of precision and recall, and the overall correctness of the model, respectively.

To maintain patient safety, several safeguards were implemented into the model. Specifically, only the messaging pools for participating physicians actively monitored by the model as part of the study were incorporated as potential routing destinations. Moreover, if the model’s confidence in a message’s classification was low, the model would revert to a default status and the message was forwarded to a default nurse triage pool.

### Model Evaluation and Statistical Analyses

To evaluate model performance, we compared the real-world effects of the model in the routed (i.e., intervention) message group with the unrouted control group. The primary endpoints of interest were: the time from patient message origination to initial message interaction by a member of the healthcare staff, 2) the time from patient message origination to query resolution, and 3) the total number of message interactions by healthcare staff. Secondary outcome measures included the precision, recall, F1 scores, and accuracy of message labeling by the model between the two groups.

We used system-generated identifiers to group individual messages into larger distinct conversations between patients and staff. Message interactions were defined as any reads, forwards, and replies performed by healthcare staff. Missing data were managed by calculating the percentage of missing observations for each of the primary outcomes and, based on the observation that the overall amount was low (<10% for each group), we performed pairwise deletions of any missing observations to ensure consistency and minimize potential bias. Outliers were identified and removed from both datasets using the Z-score (using a threshold of 3) and IQR (using a multiplier of 1.5) methods.

Based on a two-tailed alpha of 0.05, we estimated that for a relatively small effect size (Cohen’s d = 0.2), a sample size of 393 unique conversations per group would provide 80% power to reject the null hypothesis of no difference in the primary outcomes between the two groups. For a medium effect size (d = 0.5), a sample size of approximately 64 conversations per group would be sufficient. For a large effect size (d = 0.8), only about 26 conversations per group would be needed.

Comparative analyses were performed using Wilcoxon rank sum tests for non-normally distributed data. Scatter and box plots were used to assess homogeneity of variances, symmetry, and normality. Categorical data were summarized as numbers and percentages, and continuous data were summarized as means with standard deviations (SD) or medians with interquartile ranges (IQR), as appropriate. All tests were performed using Python (v3.11.5).

## RESULTS

### Patient Population and Message Characteristics

The model prospectively identified and routed a total of 469 unique conversations from adult patients between September 16 and 31, 2022. An observational cohort of 402 unrouted conversations was generated in parallel from the same period. Baseline characteristics of the patient population and message characteristics for both groups of messages are shown in **Table 1**. The median (IQR) age for the intervention group was 64.2 (18.3) years, and 63% were female.

**Table 1.** Characteristics of the Patient Population and Conversations.

| Characteristic | Intervention Group<br>(N=445) | Control Group<br>(N=391) |
| --- | --- | --- |
|  | Median (IQR) or N (%) |  |
| **Patient Demographics** |  |  |
| Age, years | 64 (18) | 63 (21) |
| Age range, years | 22—98 | 19—97 |
| Female | 291 (63) | 254 (63) |
| Race or ethnicity |  |  |
| White | 306 (66) | 255 (63) |
| Black | 115 (25) | 104 (26) |
| Asian | 22 (5) | 24 (6) |
| Other<sup>1</sup> | 20 (4) | 19 (5) |
| **Conversations** |  |  |
| Total Number of Completed Conversations<sup>2</sup> | 469 | 402 |
| Messages Per Conversation | 3 (3) | 5 (5) |
| Conversation Labels |  |  |
| Urgent | 45 (10) | 62 (15) |
| Clinician | 261 (56) | 229 (57) |
| Refill | 54 (12) | 19 (5) |
| Schedule | 63 (13) | 62 (15) |
| Form | 45 (10) | 31 (8) |

1. Includes American Indian/Alaska Native, Native Hawaiian/Pacific Islander, multiple, or those declining to answer. 2. Represents the total number of unique conversations

### Primary Outcomes

A comparative analysis of the interaction metrics between the intervention and control groups is summarized in **Table 2**. Overall, healthcare staff in the intervention group exhibited a 40% reduction in the median number of total message interactions, with significant reductions in both the time to first interaction (82.8%) and the time to conversation completion (84.4%), compared to the control group. Healthcare staff in the intervention group had a median (IQR) time to first interaction of 0.2 (3.6) hours and a time to conversation completion of 4.2 (21.6) hours, compared to 1.2 (13.3) and 26.7 (76.4) hours, respectively, compared to the control group (median difference [MD] in time to first interaction, −1.0 hours; P<0.001; time to conversation completion, −22.5 hours; P<0.001). The median (IQR) number of message interactions per conversation was also significantly lower in the intervention group at 3.0 (3.0) compared to 5.0 (5.0) total interactions in the control group (MD −2.0; P<0.001). **Figure 2** illustrates the distributions in the primary outcomes between the intervention and control groups.

**Figure 2.**
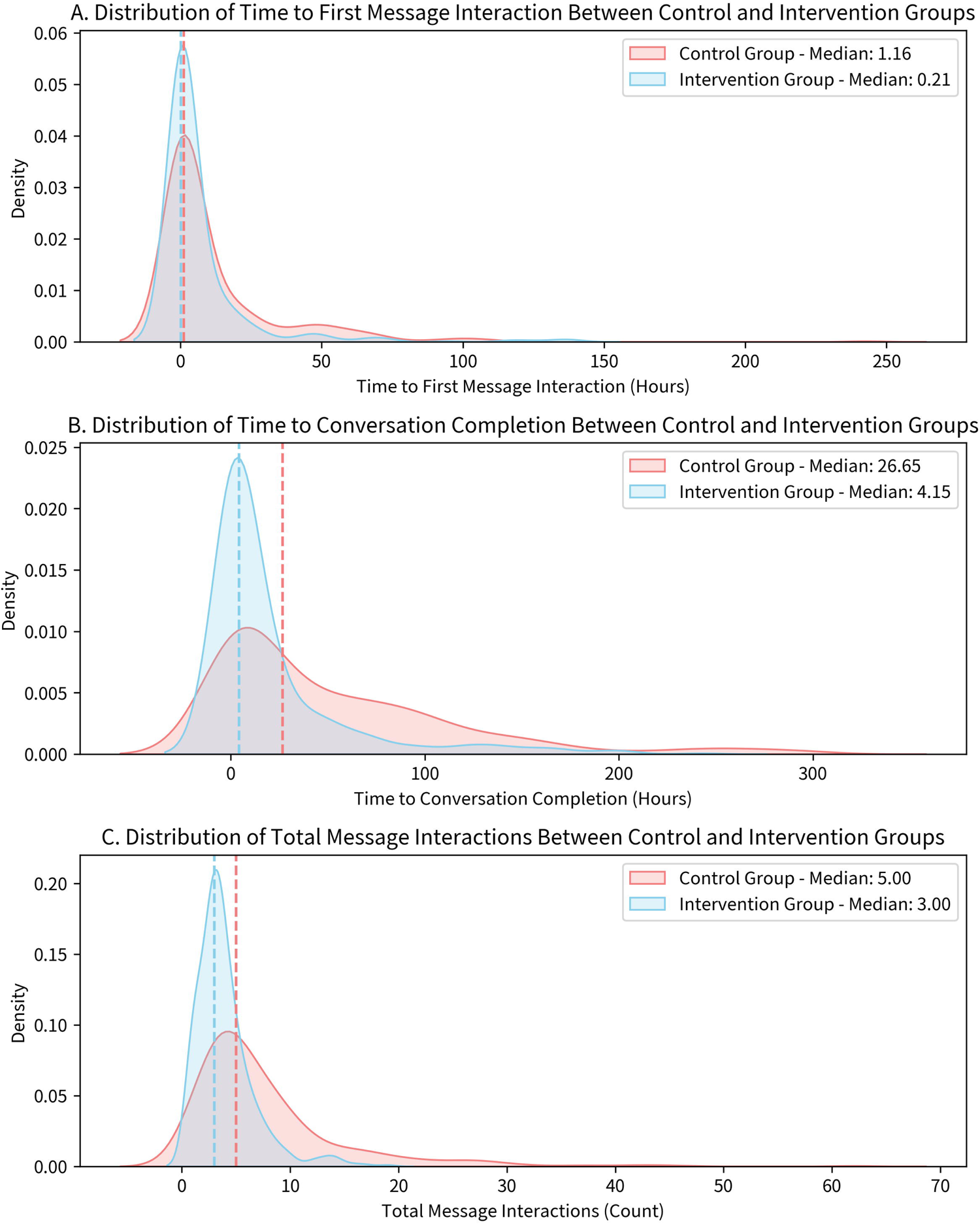
Distributions of the time to first message interaction (A), time to conversation completion (B), and total message interactions (C) between the intervention (blue) and control (red) groups.

**Table 2.** Comparison of Prospective Model Performance After Deployment.

| **Outcome** | **Intervention Group**<br>**(N=469)<sup>1</sup>** | **Control Group**<br>**(N=402)<sup>1</sup>** | **Median Difference** | **RRR (%)** | **P Value** |
| --- | --- | --- | --- | --- | --- |
| **Time to first message interaction** (median [IQR]) – hours | 0.2 [3.6] | 1.2 [13.3] | -1.0 | 83.3 | <0.001 |
| **Time to conversation completion** (median [IQR]) – hours | 4.2 [21.6] | 26.7 [76.4] | -22.5 | 84.3 | <0.001 |
| **Number of total message interactions** (median [IQR]) | 3.0 [3.0] | 5.0 [5.0] | -2.0 | 40.0 | <0.001 |

1. Represents the number of total unique conversations.

### Secondary Outcomes and Model Classification Performance

The model achieved consistently high performance for accurately predicting message classes compared to expert labeling by study clinicians (**Table 3**). The highest performance was seen for the ‘Schedule’ and ‘Refill’ classes with F1 scores of 98.1% and 99.0% and accuracies of 99.4% and 99.7%, respectively, with a total model accuracy of 97.8%. In **Figure 3**, the model demonstrated a high degree of accuracy using a correlation heatmap for analysis, with most instances correctly classified, as evident by the predominant values along the diagonal. The instances of misclassification were minimal, consistent with a high degree of discriminatory ability between different classes. There were no instances where the model defaulted back to a default mode due to low confidence in classifying an individual message.

**Table 3.** Model Performance by Message Class.

| Class | Precision | Recall | F1 Score | Accuracy |
| --- | --- | --- | --- | --- |
| Urgent | 98.1% | 95.0% | 96.5% | 98.7% |
| Clinician | 97.6% | 98.1% | 97.9% | 98.5% |
| Refill | 98.0% | 100.0% | 99.0% | 99.7% |
| Schedule | 99.2% | 96.9% | 98.1% | 99.4% |
| Form | 96.9% | 99.6% | 98.2% | 99.5% |

**Figure 3.**
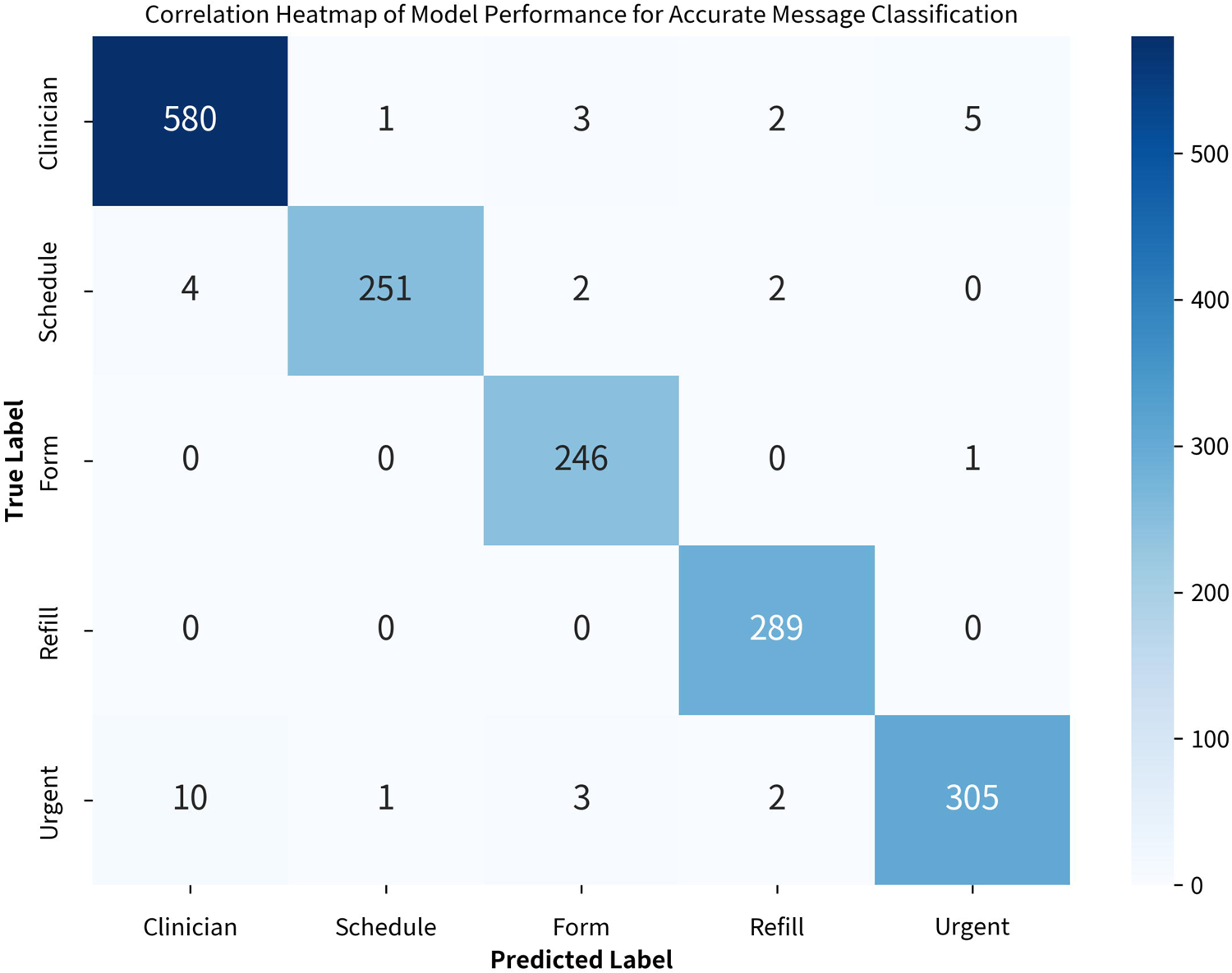
Correlation Heatmap of Model Performance for Accurate Message Classification. A confusion matrix, represented as a heatmap, illustrating the number of correct and incorrect predictions made by the model for each class. Each row of the matrix represents the true class based on expert clinician labeling, while each column represents the predicted class generated by the model. The diagonal elements represent the number of correct predictions for each class, while the off-diagonal elements in purple indicate instances where the model misclassified. A combination of high values in the diagonal elements and low values in the off-diagonal elements signifies more accurate predictions.

## DISCUSSION

Real-time message classification using a novel NLP model with intelligent rerouting was associated with significantly shorter message response and resolution times, as well as significant reductions in overall message burden for healthcare staff, when compared to a cohort of unrouted messages from the same period. On a sensitivity analysis, the effects of implementing the NLP remained consistent across most staff roles except for non-physician practitioners who exhibited a non-significant trend toward improvement with implementation of NLP message routing, which is potentially explained by their lower number of messages at baseline. The model also demonstrated high accuracy for correctly predicting each message class when compared to expert reviewers, with overall accuracy for message classification exceeding 97%.

In this study, we applied a deep learning NLP model to prospectively classify and route patient messages in real time. We used an open-source large language model (BERT) which offered several advantages over other available large language models. As a bi-directional transformer model, BERT was designed to simultaneously read text in both directions (i.e., it considered both the left and right context when making predictions) which makes it better suited for sentiment analysis and natural language understanding tasks. Compared to “uni-directional” models such as Generative Pre-trained Transformers (GPT), bi-directional models provide enhanced contextual interpretation with a lower risk of model “hallucination” – a phenomenon when an AI model generates incorrect predictions but presents these as fact – since they are simply placing the messages into predetermined categories. Additional benefits of BERT include increased computational efficiency from a reduced reliance on power-intensive graphical processing units to train and deploy NLP models into practice – resulting in potential cost savings.

Our findings are unique in implementing an advanced NLP classifier to the real-world messaging data from the EHR at multiple locations in a large healthcare system. Specific strengths of our study include the real-world setting and pragmatic trial design which yielded a robust sample size with adequate power to detect even small sample sizes, the consistency of the findings on the stratified analyses based on provider specialty, and the use of physician informaticists (Dr. Anderson) who understood both the clinical context as well as the implication on the model accuracy which facilitated creation of a labeling heuristic which allowed for classification with minimal error.

Limitations of this study include the absence of a randomized comparator group. Although we attempted to reduce the risk of bias with a comparator group through random sampling (but drawn from the same time period and matched for provider specialty) and the patient characteristics between the treatment and control groups appear similar in key demographic factors, this does not guarantee the removal of residual confounding. The study was also limited to volunteer physicians and staff at a single healthcare institution in Atlanta, GA, which could affect generalizability.

## Conclusions

In this real-world feasibility study, implementation of an NLP classifier in the EHR for intelligent message rerouting led to decreased message response times and reduced the overall messaging burden among healthcare staff, as compared to unrouted messages from the same period. These findings have the potential to significantly enhance operational efficiency within healthcare organizations and reduce administrative burden and EHR-related burnout among healthcare workers. Further validation and prospective randomized trials are needed to better understand the effects on patient outcomes and quality of care, specifically any reductions in medical errors that may result.

## Data Availability

The participants of this study have not provided written consent for their data to be shared publicly, therefore supporting data is not available due to the sensitive nature of the research

## Notes

### Competing Interest Statement

All authors have completed the ICMJE uniform disclosure form at www.icmje.org/coi_disclosure.pdf and declare: no support from any organisation for the submitted work; AH and AMH have received in-kind research support from Switchboard MD; MZ and ADC are consultants for Switchboard MD; BA is an employee of Switchboard MD; no other relationships or activities that could appear to have influenced the submitted work.

### Funding Statement

The study was funded by Switchboard MD (Atlanta, GA), an artificial intelligence and data science company, and performed in partnership with Emory University.

### Author Declarations

This was a prospective, open-label, pragmatic, feasibility study conducted at Emory Healthcare, an integrated health system in Atlanta, Georgia. The study was deemed exempt from human subjects research by the institutional review board at Emory University and received a waiver of informed consent.

